# Isolating Effects of Medicare Code Slot Expansion on Longitudinal Risk Assessment

**DOI:** 10.1101/19010074

**Authors:** Rohan Khera, Faseeha Altaf, Yongfei Wang, Susannah M. Bernheim, Zhenqiu Lin, Sharon-Lise T. Normand, Harlan M. Krumholz

**Affiliations:** Division of Cardiology, University of Texas Southwestern Medical Center, Dallas, TX; Center for Outcomes Research and Evaluation, Yale-New Haven Hospital, New Haven, CT; Section of Cardiovascular Medicine, Department of Internal Medicine, Yale School of Medicine, New Haven, CT; Section of General Medicine, Department of Internal Medicine, Yale School of Medicine, New Haven, CT; Department of Biostatistics, T.H. Chan School of Public Health, Harvard University, Boston, MA; Department of Health Care Policy, Harvard Medical School, Boston, MA; Department of Health Policy and Management, Yale School of Public Health, New Haven, CT

## Abstract

**Background:** The evaluation of trends in patient outcomes requires adjustment for the changes in case-mix over time and, thus, could be influenced by the expansion of code slots on inpatient claims that occurred in January 2011. We tested the hypothesis that the changes in code slots caused an artifactual increase in the case mix over time compared with a strategy that restricted inpatient codes to the same number of slots over time, excluding consideration of codes beyond the first 9 after the expansion.

**Methods:** In Medicare claims over a 5-year period spanning the inpatient code slot expansion (2008-2012), we created cohorts of hospitalizations for heart failure (HF), acute myocardial infarction (AMI) and pneumonia, common hospitalization conditions included in federal policies. We obtained information on risk factors for 30-day post-discharge mortality or readmission for each condition from inpatient facility Medicare claims, outpatient facility claims and professional (or carrier) claims. We evaluated the effects of additional codes captured from the expanded slots in inpatient claims on the number of risk factors or model covariates, overall and based on their contribution to the risk of mortality or readmission. We modelled the effects of code expansion on risk-assessment using an interrupted time series framework.

**Results:** There were 2,102,509 eligible hospitalizations for HF, 872,734 for AMI and 1,824,079 for pneumonia. The average number of risk factors increased across all covariate selection strategies. There was a larger increase in monthly average covariate count that included all codes at the time of the code slot increase relative to a strategy that consistently used only 9 inpatient codes (level change in interrupted time series model, 0.9% [95% CI 0.7% to 1.1%] in HF, 0.6% [0.5% to 0.7%] for AMI and 0.6% [0.4% to 0.8%] for pneumonia). Using both inpatient and outpatient/carrier codes for assessing risk factors, there was a smaller difference between strategies using 9 inpatient codes, compared with all inpatient claims (relative excess increase in covariates by 0.6% [0.4% to 0.8%] in HF, 0.4% [0.3% to 0.5%] for AMI, and 0.3% [0.1% to 0.6%] in pneumonia). However, the additional codes were limited to covariates with small contributions to the risk-adjustment models for mortality, without a significant inflection in measured risk of mortality across code expansion (P>0.05 in interrupted time-series models). Measured readmission risk increased with using only inpatient claims for risk assessment, but not if all outpatient and carrier claims were also used (P>0.05 in interrupted time-series models).

**Conclusions:** The expansion of inpatient code slots did not meaningfully affect the measurement of the risk of mortality or readmission, especially if comprehensive inpatient and outpatient claims are used, because the additional covariates only included conditions with a modest influence on risk adjusted models. The use of all versus limited codes after the code slot expansion has a minimal effect on evaluating trends in these conditions.

## BACKGROUND

The evaluation of trends in patient outcomes requires adjustment for the changes in case mix over time. Changes in Medicare coding rules, with the expansion of code slots on inpatient claims that occurred in January 2011, could influence risk adjustment independent of any changes in the patient population and lead to false inferences about the effect of time. This issue can be particularly important when evaluating the effect of policy changes around the time of the code slot expansion, such as occurred with the Hospital Readmission Reduction Program (HRRP). ^1-3^

Studies that have evaluated the effect of different methodological approaches to these changes have been limited. ^2,3^They have focused on claims derived from limited sources, such as inpatient claims,^1^or inpatient and limited outpatient claims,^2^ as opposed to the use of more comprehensive claims data, as is used in the CMS measures, which may blunt the impact of the change. Also, at least one study only evaluated changes in frequency of a few select comorbidities rather than the effect on risk or risk-adjusted trends.^1^There is a need to determine how different methodological approaches to managing the changes in the coding rules affects characterization of risk of readmission and mortality.

Accordingly, we tested the hypothesis that the changes in code slots caused an artifactual increase in the case mix over time compared with a strategy that restricted inpatient codes to the same number of slots over time, excluding consideration of coders beyond the first 9 after the expansion. We used the entire complement of Medicare claims from inpatient and outpatient care settings, and focused on the 3 conditions that are frequently targeted in Federal policies to define study cohorts. These 3 conditions - heart failure, acute myocardial infarction (AMI), and pneumonia – have also been a part of the HRRP since its inception. Although we focus on conditions included in the HRRP, this issue also has importance for any assessments of change in practices and outcomes conducted over this period.

## METHODS

### Overview

We tested different strategies for the number of inpatient codes use and the data sources used on the change, at the time of the coding increase, on the frequency of comorbidities and on the risk of readmission or mortality.

### Data Sources

We used the 100% Medicare Standard Analytic Files for the years 2008 through 2012 that included all claims submitted for fee-for-service Medicare beneficiaries from inpatient, outpatient, and physician professional claims.^3^Inpatient claims represent Medicare claims submitted by hospitals for inpatient hospitalizations, including both diagnoses and procedures that were part of the treatment during the hospitalization.^4^Research studies frequently use the inpatient complement of claims.^5^Outpatient claims represent facility claims that correspond to clinic visits, observation stays, emergency department visits, and outpatient rehabilitation center visits.^6^Certain studies leverage these outpatient claims as well.^2,7^The physician provider or carrier claims, which numerically represent the largest set of claims, are submitted by physicians and other clinical providers for professional clinical services during either inpatient or outpatient visits, including services where facility charges are not submitted.^8^The availability of these data is limited.^8^The measures for the Centers of Medicare and Medicaid Services (CMS) use all these sources of data to define comorbidities that are used in risk-adjustment models. In the current study, the term “inpatient claims” refers to institutional claims for hospitalizations, and “outpatient claims” refer to all outpatient facility claims and professional claims across all settings.

### Study Population

We included all hospitalizations for patients discharge alive after an inpatient hospital admission with one of the 3 conditions that have always been included in the Hospital Readmissions Reduction Program – heart failure, acute myocardial infarction (AMI), or pneumonia. Index hospitalizations for these conditions were defined by a principle admission diagnosis for the respective conditions on inpatient claims. The definitions for these hospitalizations are consistent with those used in the CMS measures to the define these conditions,^9,10^ and with studies that have used Medicare data to evaluate the HRRP (**eFigures 1-3**).^11-13^

### Covariate selection strategies

Covariates for risk adjustment were those used in the CMS 30-day readmission and mortality based on claims submitted in the 12-month period preceding the index hospitalization event. We identified 4 covariate selection strategies based on whether studies used only inpatient claims or both inpatient and comprehensive outpatient claims to identify comorbid health conditions, and how they processed inpatient claims after code slots increased in 2011. These strategies identified covariates using (1) only inpatient claims, using 9 secondary diagnosis codes and 6 procedure codes even when more codes were available, (2) only inpatient claims, using all available codes after the expansion of codes, (3) a fixed number of inpatient claims (as in 1, above), but supplemented with comprehensive outpatient claims, and (4) all inpatient claims (as in 2, above) after the code expansion, along with comprehensive outpatient claims.

### Statistical analyses

First, we examined trends in the number of covariates for each month in the 5-year period spanning the transition based on each of the 4 covariate selection strategies. Next, to identify the relative contribution of each covariate in the adjustment models for readmission and mortality, using data in the year 2008, for each of the 3 HRRP conditions, we constructed logistic regression models with mortality and readmission as outcomes, and the set of variables included in the respective risk-adjustment models of the CMS from inpatient and outpatient claims before any expansion of the code slots. The regression coefficients for the variables in these models represent the contribution of each covariate to the risk of readmission and mortality among patients. Next, we identified covariates that were associated with the highest risk of death and readmission across the 3 HRRP conditions based on their regression coefficients relative to others. We evaluated temporal trends in the capture of these conditions for each of the 4 covariate selection strategies. Finally, we assessed how the overall risk of mortality and readmission, based on the model changed across the expansion of the codes.

All other analyses were performed using SAS, version 9.4 (Cary, NC), and Stata 14 (College Station, TX). The level of significance was set at 0.05. The study was reviewed by the Yale University Institutional Review Board and deemed exempt from informed consent due to the deidentified data.

## RESULTS

There were 2,102,509 eligible hospitalizations for HF, 872,734 for AMI and 1,824,079 for pneumonia over the study period (**eFigures 1-3**).

### Change in Number of Risk Factors

The average number of risk factors increased across all covariate selection strategies. There was a relative increase in the average number of covariates in the strategies that used all inpatient codes after expansion the code slots for all 3 conditions (**Figure 1, and eFigure 4**). There was a relative excess increase in mean covariates count at code announcement of 0.9% (level change in interrupted time series model, 95% CI 0.7% to 1.1%), in heart failure, 0.6% (0.5% to 0.7%) for AMI and 0.6% (0.4% to 0.8%) for pneumonia, relative to a strategy that consistently used only 9 inpatient codes (**Table 1**).

**Table 1.** Heart Failure, Acute Myocardial Infarction, and Pneumonia: Number of Risk Factors, Mortality and Readmission Risk Scores in Post-Expansion Period.

|  | Relative level change in post-expansion period |  | Relative slope change in post-expansion period |  |
| --- | --- | --- | --- | --- |
|  | Absolute level change, % (95% CI) | P value | Absolute level change, % (95% CI) | P value |
| **HEART FAILURE** |  |  |  |  |
| **Inpatient codes only** |  |  |  |  |
| Number of risk factors | 0.922 (0.724, 1.121) | <0.001 | 0.027 (0.012, 0.043) | 0.001 |
| Mortality risk score | -0.028 (-0.095, 0.039) | 0.41 | 0.000 (-0.004, 0.004) | 0.87 |
| Readmission risk score | 0.049 (0.022, 0.076) | <0.001 | 0.002 (-0.000, 0.004) | 0.050 |
| **Inpatient and outpatient codes** |  |  |  |  |
| Number of risk factors | 0.568 (0.354, 0.781) | <0.001 | 0.014 (-0.001, 0.030) | 0.07 |
| Mortality risk score | -0.024 (-0.089, 0.042) | 0.48 | 0.000 (-0.004, 0.004) | 0.92 |
| Readmission risk score | 0.025 (-0.003, 0.052) | 0.08 | 0.001 (-0.001, 0.003) | 0.31 |
| **ACUTE MYOCARDIAL INFARCTION** |  |  |  |  |
| **Inpatient codes only** |  |  |  |  |
| Number of risk factors | 0.602 (0.525, 0.678) | <0.001 | 0.009 (0.004, 0.013) | <0.001 |
| Mortality risk score | -0.031 (-0.130, 0.068) | 0.54 | -0.000 (-0.006, 0.006) | 0.97 |
| Readmission risk score | 0.037 (-0.009, 0.082) | 0.11 | 0.001 (-0.002, 0.003) | 0.68 |
| **Inpatient and outpatient codes** |  |  |  |  |
| Number of risk factors | 0.410 (0.339, 0.481) | <0.001 | 0.005 (0.001, 0.009) | 0.021 |
| Mortality risk score | -0.029 (-0.128, 0.070) | 0.56 | 0.000 (-0.006, 0.006) | 0.94 |
| Readmission risk score | 0.018 (-0.028, 0.064) | 0.44 | 0.000 (-0.003, 0.003) | 0.89 |
| **PNEUMONIA** |  |  |  |  |
| **Inpatient codes only** |  |  |  |  |
| Number of risk factors | 0.582 (0.367, 0.796) | <0.001 | 0.017 (-0.001, 0.035) | 0.07 |
| Mortality risk score | -0.005 (-0.079, 0.069) | 0.89 | 0.000 (-0.004, 0.005) | 0.84 |
| Readmission risk score | 0.038 (0.005, 0.071) | 0.03 | 0.001 (-0.001, 0.004) | 0.25 |
| **Inpatient and outpatient codes** |  |  |  |  |
| Number of risk factors | 0.342 (0.067, 0.616) | 0.02 | 0.008 (-0.013, 0.030) | 0.45 |
| Mortality risk score | -0.005 (-0.081, 0.072) | 0.91 | 0.000 (-0.004, 0.005) | 0.88 |
| Readmission risk score | 0.020 (-0.017, 0.057) | 0.29 | 0.001 (-0.002, 0.003) | 0.61 |

**Figure 1:**
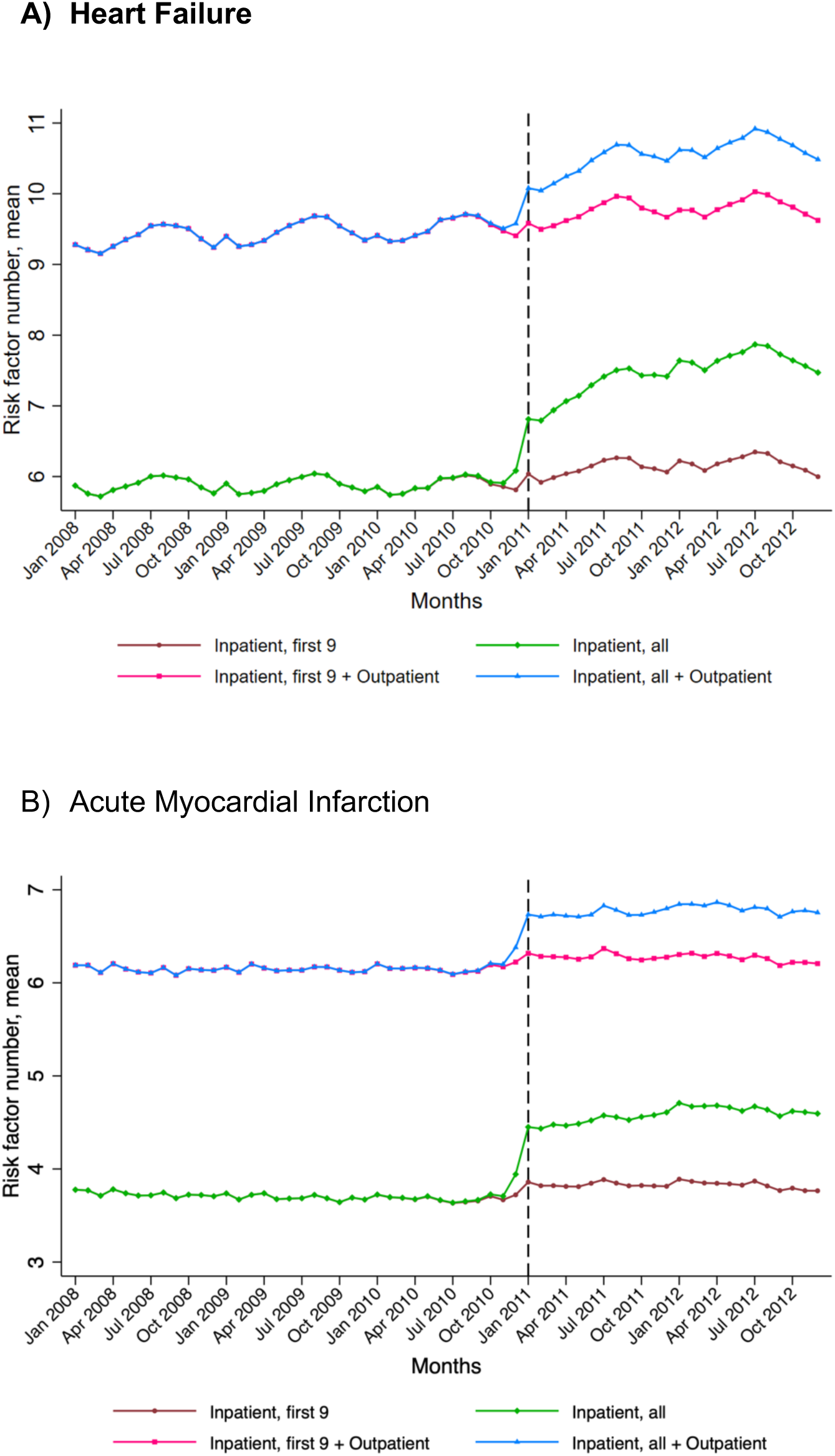
Trends in risk factor number for heart failure and Acute myocardial infarction.

In the strategy that used both inpatient and outpatient codes for assessing risk factors, there was a smaller but significant relative increase in the average number of codes when all codes were used after inpatient code slot expansion compared with a strategy that combined the used a fixed number of inpatient codes with all outpatient codes (**Figure 1**). There was a 0.6% (0.4% to 0.8%) increase in covariates for heart failure, 0.4% (0.3% to 0.5%) for AMI, and 0.3% (0.1% to 0.6%) in pneumonia. There was a small but significant continued relative increase in covariates captured in strategies using all codes in AMI, but not for heart failure or pneumonia, measured as difference in slopes after accounting for the level change after code expansion (**Table 1**).

### Changes in Covariates by Risk Contribution

The change in captured risk factors is not consistent across covariates, with a significant increase in certain covariates, e.g. history of coronary artery bypass grafting, but not others, such as metastatic cancer and malnutrition. After ordering covariates by their contribution to the risk of mortality based on the magnitude and direction of the regression coefficient in risk-adjustment models, the top 10 covariates experienced changes in covariate frequency that were similar in magnitude and direction regardless of whether the first 9 or the complete inpatient claims were used after expansion of code slots, and regardless of the use of outpatient codes (**Table 2, Figure 2, eFigures 5-6**). Notably, substantial discordance in relative trends of covariate were mainly observed for covariates which conferred the lowest risk of mortality in risk-adjustment models (**Table 2, Figure 2, eFigures 5-6**). In 30-day readmission models, however, some of the top 10 covariates demonstrated a relative increase with the use of all inpatient codes after code slot expansion, relative to strategies that used a fixed number of codes (**eFigures 7-8**).

**Table 2:** Covariate importance to heart failure mortality model and average annual change in covariate frequency. Covariates are ranked in descending order of their regression coefficient in the heart failure model for post-discharge 30-day mortality. Average annual absolute percentage change in the covariate frequency based on 4 covariate selection strategies during 2008-2012 are presented.

| Covariate rank | Covariate | Regression coefficient |  | Average annual change in covariate frequency |  |  |  |
| --- | --- | --- | --- | --- | --- | --- | --- |
|  |  | Estimate | S.E. | Inpatient, first 9 | Inpatient, all | Inpatient, first 9 + Outpatient | Inpatient, all + Outpatient |
| 1 | Metastatic Cancer and Acute Leukemia (CC 7) | 0.96 | 0.03 | -0.001 | -0.001 | 0.000 | 0.000 |
| 2 | Protein-Calorie Malnutrition (CC 21) | 0.60 | 0.02 | 0.057 | 0.074 | 0.063 | 0.079 |
| 3 | Decubitus ulcer or chronic skin ulcer (CC 148, 149) | 0.33 | 0.02 | -0.005 | 0.028 | 0.013 | 0.032 |
| 4 | Dementia and Senility (CC 49, 50) | 0.29 | 0.01 | 0.033 | 0.143 | 0.022 | 0.094 |
| 5 | Pneumonia (CC 111 to 113) | 0.20 | 0.01 | 0.038 | 0.041 | 0.042 | 0.043 |
| 6 | Congestive Heart failure (CC 80) | 0.18 | 0.02 | 0.042 | 0.066 | 0.015 | 0.021 |
| 7 | Severe hematological disorders (CC 44) | 0.17 | 0.03 | -0.015 | -0.007 | -0.017 | -0.013 |
| 8 | Renal Failure (CC 131) | 0.16 | 0.01 | 0.161 | 0.208 | 0.205 | 0.232 |
| 9 | Disorders of fluid/electrolyte/acid-base (CC22, 23) | 0.16 | 0.01 | 0.092 | 0.200 | 0.108 | 0.178 |
| 10 | Major psychiatric disorders (CC 54 to 56) | 0.13 | 0.02 | 0.010 | 0.040 | 0.023 | 0.044 |
| 11 | Cardio-respiratory failure and shock (CC 79) | 0.13 | 0.01 | 0.106 | 0.140 | 0.107 | 0.128 |
| 12 | Hemiplegia, Paralysis, Functional Disability (CC 67 to 69, 100 to 102, 177, 178) | 0.13 | 0.02 | 0.014 | 0.044 | 0.022 | 0.041 |
| 13 | Liver and biliary disease (CC 25 to 30) | 0.10 | 0.02 | 0.019 | 0.055 | 0.034 | 0.060 |
| 14 | Chronic Obstructive Pulmonary Disease (CC 108) | 0.10 | 0.01 | 0.014 | 0.128 | 0.017 | 0.084 |
| 15 | Fibrosis of lung and other chronic lung disorders (CC 109) | 0.08 | 0.02 | 0.001 | 0.031 | -0.038 | -0.017 |
| 16 | Drug/alcohol abuse/dependence/psychosis (CC 51 to 53) | 0.06 | 0.02 | 0.004 | 0.108 | 0.029 | 0.107 |
| 17 | Acute coronary syndrome (CC 81, 82) | 0.06 | 0.02 | -0.013 | -0.011 | -0.025 | -0.024 |
| 18 | Peptic ulcer, hemorrhage, other specified gastrointestinal disorders (CC 34) | 0.04 | 0.02 | -0.002 | 0.015 | -0.002 | 0.009 |
| 19 | Other psychiatric disorders (CC 60) | 0.03 | 0.02 | 0.035 | 0.187 | 0.073 | 0.189 |
| 20 | Valvular or Rheumatic Heart Disease (CC 86) | 0.03 | 0.01 | 0.013 | 0.273 | 0.025 | 0.179 |
| 21 | Depression (CC 58) | 0.02 | 0.02 | 0.006 | 0.209 | 0.046 | 0.197 |
| 22 | Vascular or Circulatory Disease (CC 104, 105, 106) | 0.02 | 0.01 | 0.024 | 0.177 | 0.054 | 0.110 |
| 23 | Stroke (CC 95, 96) | 0.01 | 0.02 | 0.000 | 0.000 | -0.013 | -0.013 |
| 24 | Other urinary tract disorders (CC 136) | 0.01 | 0.01 | -0.056 | 0.091 | -0.028 | 0.061 |
| 25 | Iron deficiency and other/unspecified anemias and blood disease (CC 47) | 0.00 | 0.01 | 0.101 | 0.430 | 0.096 | 0.261 |
| 26 | End-stage renal disease or dialysis (CC 129, 130) | -0.01 | 0.03 | 0.007 | 0.027 | 0.022 | 0.026 |
| 27 | Arrhythmias (CC 92, 93) | -0.02 | 0.01 | 0.042 | 0.141 | 0.075 | 0.105 |
| 28 | Cancer (CC 8 to 12) | -0.03 | 0.02 | 0.003 | 0.043 | 0.001 | 0.019 |
| 29 | Nephritis (CC132) | -0.05 | 0.04 | 0.001 | 0.035 | 0.003 | 0.033 |
| 30 | Other and unspecified heart disease (CC 94) | -0.06 | 0.01 | -0.012 | 0.100 | -0.030 | 0.038 |
| 31 | Diabetes and DM Complications (CC 15 to 20, 119, 120) | -0.10 | 0.01 | 0.043 | 0.155 | 0.046 | 0.078 |
| 32 | Other gastrointestinal disorders (CC 36) | -0.11 | 0.01 | 0.026 | 0.427 | 0.060 | 0.249 |
| 33 | Chronic atherosclerosis (CC 83, 84) | -0.17 | 0.01 | -0.048 | 0.160 | -0.035 | 0.055 |
| 34 | Asthma (CC 110) | -0.26 | 0.02 | 0.000 | 0.030 | 0.006 | 0.027 |
| 35 | Coronary Artery Bypass Graft Surgery | -0.36 | 0.02 | -0.031 | 0.239 | -0.031 | 0.239 |

**Figure 2:**
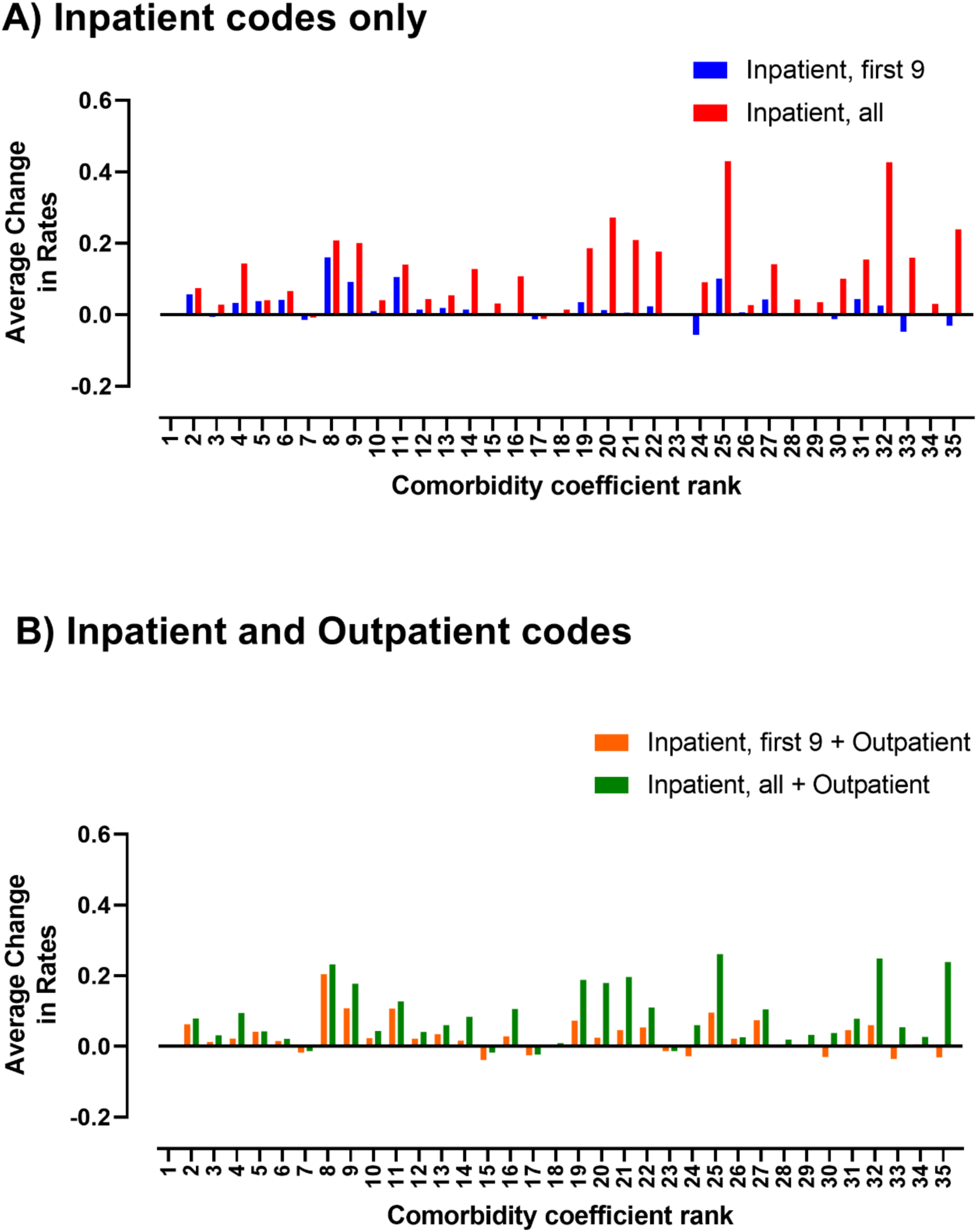
Covariate contribution to risk adjustment model for mortality and relative excess increase with code expansion in Heart Failure.

### Coded Risk of Mortality and Readmission

Changes in coded severity of illness for the mortality and readmission outcomes were sensitive to the sources of data. For mortality, across all 3 conditions, after the increase in code slots, the use of all inpatient codes was associated with a lower coded severity of illness compared with the strategy using only 9 codes consistently (**Figure 3**). There were no inflections in the coded severity with the use of the full complement of inpatient and outpatient codes, compared with first 9 inpatient with all outpatient claims (P for level change in interrupted time series models > 0.05 for all, **Table 1**).

**Figure 3:**
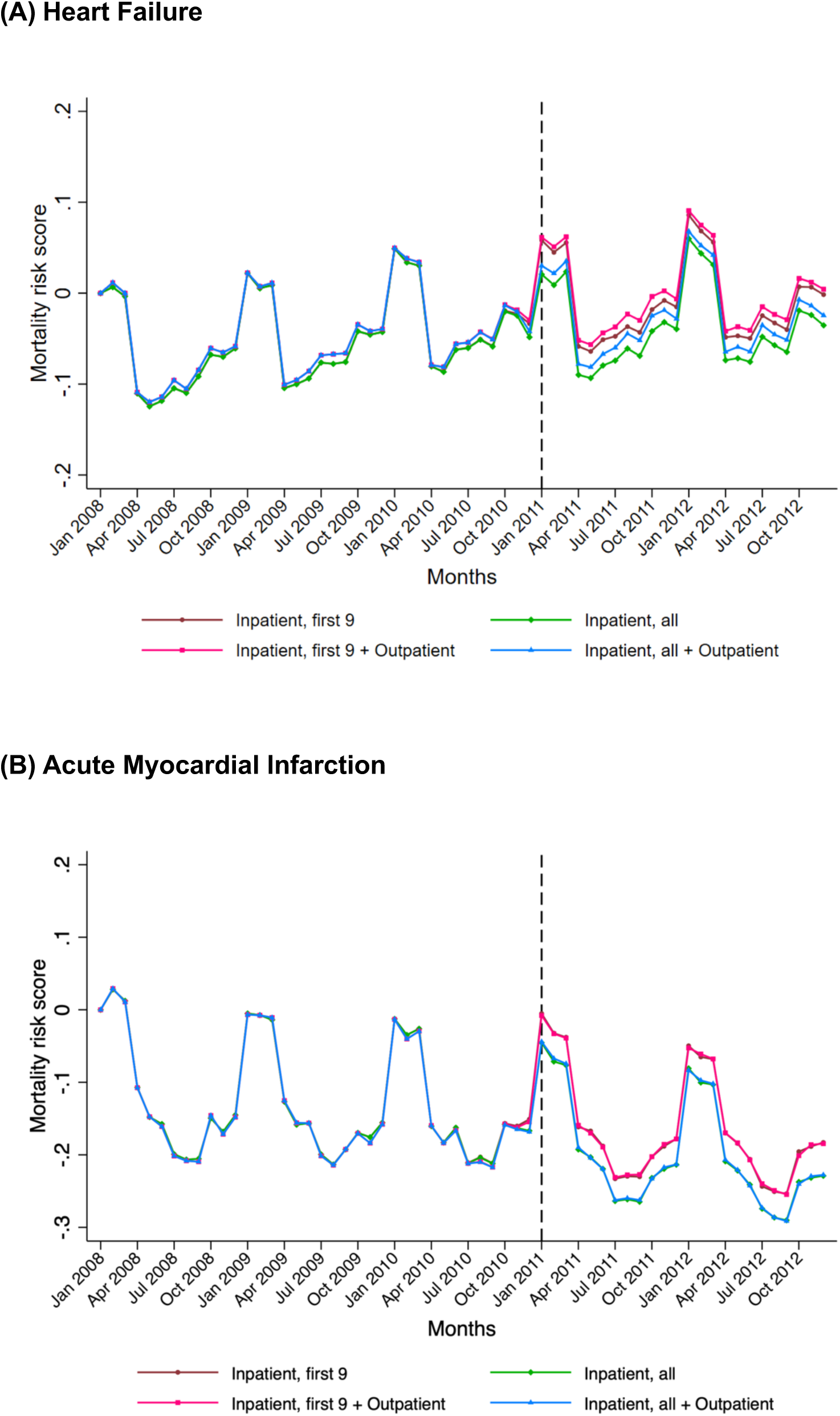
Monthly Risk Score for 30-day Post-discharge Mortality.

**Figure 4:**
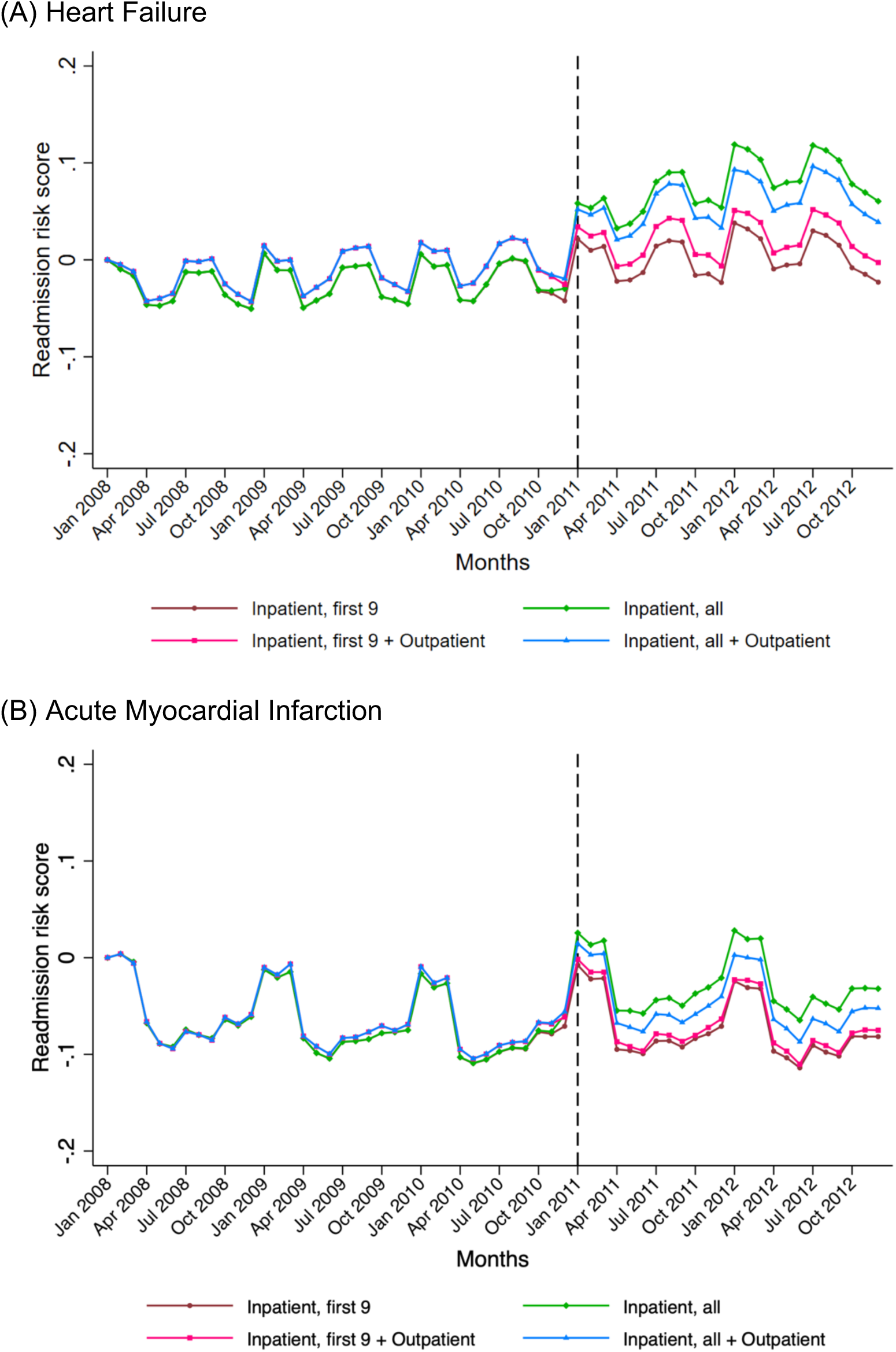
Monthly Risk Score for 30-day Readmission.

The readmission risk score did not increase if both inpatient and outpatient data were used for identifying covariates regardless of whether all or only 9 inpatient codes were used after increase in the number of code slots (**Table 1**). In contrast, if only inpatient codes were used as the source of the data, there was a relative excess increase in the readmission risk with code expansion when using all inpatient codes, compared with only 9 inpatient codes.

## DISCUSSION

We found that the use of comprehensive inpatient and outpatient codes, with a 12 month look back, as is done with the CMS measures, blunts the effect of the code expansion on increasing the comorbidity prevalence. Moreover, the additional covariates identified with the expanded code slots did not represent conditions that conferred substantial risk of mortality or readmission. The measured risk of mortality based on either coding strategy was, therefore, unchanged regardless of the number of codes used after the expansion. The risk of readmission also did not significantly increase with code expansion if outpatient codes were used in addition to the inpatient codes.

The expansion of inpatient code slots occurred during a period with several health policy interventions under the Affordable Care Act.^14^ The evaluation of these policies has relied on temporal trends, which use covariates identified from claims to account for changes in patient characteristics over time. The expansion of codes, and how different evaluations of policy handled these changes, have specifically been suggested to overestimate the effects of the national program to reduce readmission.^2^ However, the study did not use the entire complement of outpatient claim codes. Our assessment of readmission risk assessed using comprehensive outpatient codes in addition to the inpatient codes did not find any suggestion for a significant artifactual increase in readmission risk with using all available codes after the code slots increased. This effect was limited to assessments of risk that focused only on inpatient claims. Other studies that have evaluated the effect of the Hospital Readmissions Reduction Program and reached different conclusions also differ in their use of inpatient vs inpatient and outpatient data, as well as the use of 9 inpatient or all available codes for risk adjustment.^3,5^ Our findings of no inflections in mortality risk with either coding strategy suggests that the differences between these studies do not arise from the data source or their strategy of handling covariates for risk adjustment.

The design of the CMS measures, which included inpatient and outpatient codes over the prior year, was intended to comprehensively characterize comorbidity burden and outcome risk from multiple sources. Although the intent to expand codes was not known when we developed the original outcomes measures,^11-13,15-17^ there was a concern that over-reliance on the index hospitalization, or hospitalizations alone, could yield to gaming strategies. Including comprehensive data made it more difficult for coding strategies by institutions to influence the risk adjustment. Our study indicates that this strategy also mitigated the effect of the coding rule change.

The study has certain limitations. We are unable to identify the coding strategies that best correspond to the actual patient severity over time and are only limited to comparing relative changes across different coding strategies. Moreover, the study does not account for heterogeneity in coding changes across US hospitals and has mainly focused on the evaluation of overall trends. The analyses focus on the period around the code expansion, using a fixed code strategy as the control. However, we are unable to quantify the effect of the code expansion on the quality of this control group in the years beyond the analysis period.

In conclusion, the expansion of inpatient code slots led to the identification of a larger number of covariates included in risk-adjusted models across the transition. However, the additional covariates did not meaningfully affect the measurement of the risk of mortality or readmission, especially if both inpatient and outpatient claims were used, as the additional covariates only included conditions without a significant influence on risk adjusted models. The hypothesis that strategies of covariate selection to risk-adjust, therefore, does not explain temporal trends in readmission and mortality.

## Data Availability

The data are proprietary of the CMS but can be obtained from them directly under a data use agreement directly with the CMS,

## ACKNOWLEDGMENT

### Funding

Dr. Khera is supported by the National Center for Advancing Translational Sciences (UL1TR001105) of the National Institutes of Health. The funder had no role in the design and conduct of the study; collection, management, analysis, and interpretation of the data; preparation, review, or approval of the manuscript; and decision to submit the manuscript for publication.

### Disclosures

Dr. Krumholz is a recipient of a research grant, through Yale, from Medtronic and Johnson & Johnson (Janssen) to develop methods of clinical trial data sharing; was a recipient of a research grant, through Yale, from Medtronic and the Food and Drug Administration to develop methods for post-market surveillance of medical devices; was a recipient of a research agreement, through Yale, from the Shenzhen Center for Health Information for work to advance intelligent disease prevention and health promotion, and collaborates with the National Center for Cardiovascular Diseases in Beijing; received payment from the Arnold & Porter Law Firm for work related to the Sanofi clopidogrel litigation and from the Ben C. Martin Law Firm for work related to the Cook IVC filter litigation; chairs a cardiac scientific advisory board for UnitedHealth; is a participant/participant representative of the IBM Watson Health Life Sciences Board; is a member of the Advisory Board for Element Science, the Physician Advisory Board for Aetna, and the Advisory Board for Facebook; and is the founder of Hugo, a personal health information platform. Drs. Krumholz, Bernheim, and Lin, and Mr. Wang work under contract with the Centers for Medicare & Medicaid Services to develop and maintain performance measures that are publicly reported. The other authors report no potential conflicts of interest.

The study was conceived and conducted by the authors, and the Centers for Medicare & Medicaid Services played no role in its design and conduct; collection, management, analysis, and interpretation of the data; and preparation, review, or approval of the manuscript.

